# Spatially Context-Aware Transformers Facilitate Modeling-Based Anomaly Detection of Subtle Lesions in Brain MRI Images

**DOI:** 10.64898/2026.09.14.26363006

**Authors:** Johannes Schwarz, Lena Will, Jörg Wellmer, Axel Mosig

## Abstract

The detection of small and subtle lesions in high-resolution 3D volumes is a highly relevant, yet far from solved task in biomedical imaging. We here address a specific task in detecting certain types of epileptogenic lesions through our novel semi-supervised *spatially context aware transformer* (SpyCAT) approach to anomaly detection. SpyCAT is modeling-based in the sense that it builds on specific assumptions that constitute what is normal and what constitues relevant deviations from normality. We explicitly use these assumptions to justify the inductive bias of our anomaly detection approach. The resulting SpyCAT system is patch-based and uses a transformer architecture to process discrete tokens obtained from a vector quantizing variational autoencoder, which produces counterfactual patches through full 3D convolutions of each patch. We evaluate our approach on the grounds of point-annotations of two subtypes of epileptogenic lesions, using validation measures that build on the *Metrics Reloaded* framework, showing that SpyCAT can reliably identify and localize the lesion types under consideration, and outperforms state-of-the-art reference methods. Our code is available at https://github.com/johannesSX/SpyCAT.

## 1. Introduction

Anomaly detection approaches have been widely investigated for the detection of brain lesions in MRI images (Simarro et al., 2020; Bowles et al., 2017; Kascenas et al., 2022; Meissen et al., 2023; Baur et al., 2021). The vast majority of these approaches have been validated on relatively large lesions, mostly brain tumors, that are correspondingly easy to detect (Luo et al., 2023; Simarro et al., 2020; Wyatt et al., 2022). However, in many cases, neurological disorders are associated with brain lesions that are much smaller and subtler than the tumors present in the data sets commonly used to assess brain anomaly detection approaches. In particular, there is a wide and heterogeneous spectrum of epilepsy associated lesions that may cover only small volumes in brain MRI images (Wellmer et al., 2013). Correspondingly, the identification and localization of such lesions via automatized anomaly detection is a much more challenging task (Spitzer et al., 2022), and the question whether subtle lesions can be detected by anomaly detection approaches must be considered open. In this contribution, we propose the *spatially context aware transformer* (SpyCAT) as a novel semi-supervised anomaly detection approach that addresses this question in the context of localizing certain subtypes of epilepsy associated brain lesions on MRI images.

As illustrated in Figure 1, our approach is based on a set of explicit assumptions which state that small and subtle lesions can be identified as anomalies that deviate in contrast to the morphological context in their non-lesional, local spatial surrounding. We translate these explicit assumptions into a transformer-based approach that operates on feature embeddings obtained from a vector quantized variational autoencoder (VQ-VAE). The feature embeddings are computed on small patches whose size reflects the expected small size of the lesions to be detected. Spatial context is provided by feature embeddings of a neighboring patch, so that a transformer network can be trained and applied to a concatenation of a repertoire of spatial context, a position encoding, and the patch of interest, in which a lesion is to be detected. This yields a transformed patch embedding, whose deviation from an expected normal, lesion-free patch can be measured against a naive patch reconstruction.

**Figure 1:**
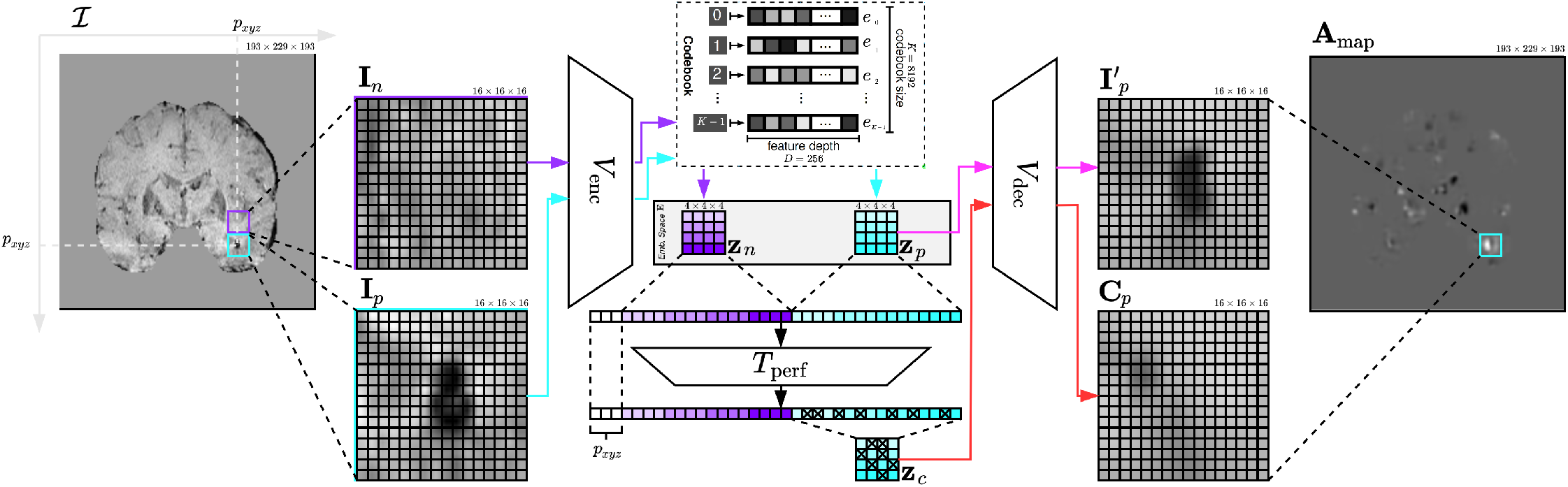
Schematic model architecture for simplicity with 2D images (sampled from coronal view of the MRI). The VQ-VAE (consists of *V*_enc_ and *V*_dec_) receives an input patch **I**_*p*_, a neighborhood patch **I**_*n*_ and generates an embedding space **z**_*p*_ and **z**_*n*_. By autoregression, the transformer *T*_perf_ creates a counterfactual (healed) embedding space **z**_*c*_, given **z**_*n*_ and a 3D position coordinate *p*, which describes where the patch is located in the MRI. Using the decoder *V*_dec_, the original embedding space **z**_*p*_ and the counterfactual embedding space **z**_*c*_ become 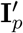 and **C**_*p*_. Subtraction between 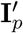 and **C**_*p*_ generates a patch anomaly map and these anomaly maps can be combined to an anomaly map **A**_*map*_ by rasterisation.

We evaluate and compare our method on two datasets against the *β*-variational autoencoder (Loizillon et al., 2024) and the f-AnoGAN (Schlegl et al., 2019; Simarro et al., 2020) as two state-of-the-art anomaly detection approaches. Our first dataset is an in-house collection of MRI dataset with two subtypes of epilepsy associated lesions (for simplicity reaons epilepsy associated or epileptogenic lesion are summarized under the acronym EL). As a second dataset, we assess detection performance on the well-established BraTS dataset for detecting brain tumors.

## 2. Related Work

Anomaly detection has been widely investigated within and beyond medical image analysis. Many state-of-the-art anomaly detection approaches have been developed outside the medical domain around the MVTec AD dataset (Bergmann et al., 2019) which consists of images of industrial parts. Numerous anomaly detection approaches have been proposed that deliver strong performance (Lee et al., 2022; Defard et al., 2020; Roth et al., 2021) on this particular datasrt. Yet, these approaches are tailored towards 2D applications and raise the question whether anomalies of non-medical objects in the MVTec dataset are set apart from normal data in the same manner as anomalies in medical image data. In the context of MRI images, anomaly detection approaches are often evaluated on the BraTS tumor dataset (Menze et al., 2015; Bakas et al., 2017, 2018; Schwarz et al., 2024) in which the anomalies cover a relatively large fraction of the image volume and are correspondingly clearly visible and can be identified as global anomalies utilizing 3D approaches (Simarro et al., 2020; Bowles et al., 2017; Iqbal et al., 2023; Luo et al., 2023). Many approaches work on the level of 2D slices, where the BR35H dataset (Hamada, 2020) is commonly used to investigate detection performance (Guo et al., 2024; Wyatt et al., 2022; Behrendt et al., 2023).

In some contexts, for example when detecting epilepsy associated lesions (Walger et al., 2023) or multiple sclerosis lesions (Carass et al., 2017), the lesions to be detected are smaller and more subtle than brain tumors. While multiple sclerosis lesion detection is often guided by longitudinal data (Carass et al., 2017), epileptogenic lesions usually need to be detected in single MRI images. For this purpose, in particular for detecting focal cortical dysplasias, supervised methods have been used more predominantely (Walger et al., 2023), while anomaly detection has so far taken no relevant role. Corresponding supervised classifiers have, for example, been proposed by Gill et al. (2021) and Spitzer et al. (2022). To find very small lesions, a focus hypothesis using EEG can be helpful and improves the result of the classifier (Gill et al., 2021).

In many of these studies, the lesion data are usually obtained from the same scanner and were recorded in the same hospital using one and the same protocol. As Bottani et al. (2023) have identified, the generalizability to clinical routine data is severely limited. A few approaches utilize collections of lesion-free reference data such as the IXI dataset^1^ to learn the patterns of *normal* dataset, and then identify deviations from those as lesional anomalies (Wyatt et al., 2022). However, these approaches rely either on anomaly detection in 2D sections or lack multicentric validation. On MRI data, anomaly detection is often carried out using supervised (Gill et al., 2021; Spitzer et al., 2022) and semi-supervised (Simarro et al., 2020; Iqbal et al., 2023; Luo et al., 2023; Marimont and Tarroni, 2020) methods. Annotation in 3D space being very time demanding, semi-supervised approaches are of high relevance, since training requires only *normal* MRIs, while annotated lesional MRIs are only required for validation.

Small and subtle lesions impose additional challenges to anomaly detection, since the small local deviations may be overshadowed by non-lesional morphological variances of a specific individual brain. This can in general be addressed by patch-based learning, where the brain image volume is decomposed into smaller patches. Recently, Behrendt et al. (2023) used this approach to reliably reconstruct normal brain MRI patches, which can be contrasted with matching patches in a given MRI to reveal lesions. In a sense, patch decomposition adds prior knowledge to anomaly detection, as the patch size is usually chosen according to the expected size of the lesions to be detected. This consideration is in line with the modeling approach suggested by Baur et al. (2021), in which the detection of abnormalities involves modeling assumptions what variances in the data are considered normal.

Recently, diffusion-based models exhibited strong performance in medical image classification (Favero et al., 2025) and related tasks such as the generation of counterfactual images (Fathi et al., 2024). Diffusion approaches that have been proposed are often implemented on 2D image data or on 2D slices extracted from 3D images (Wyatt et al., 2022; Behrendt et al., 2023). In the lack of diffusion approaches in full 3D, diffusion approaches have been established for detecting relatively large lesions that are detectable in 2D, such as lung pathologies in chest X-ray images.

## 3. Approach

When considering anomaly detection approaches for medical imaging data, it is inevitable to realize that anomalies can occur in many types: Some anomalies can be a sharply localized point divergence, while others can be diffuse morphological alterations or even global structural deviations compared to images of normal, healthy or wildtype entities. Thus, when applying anomaly detection in a particular setting, it is inevitable to raise the question what types of anomalies are relevant. In other words, one needs to constitute a notion of normality, and decide what types of deviations from normality represent relevant anomalies. In the lack of an established term for this constituting process, we refer to it as *contextualizing normality*.

Our SpyCAT algorithm contextualizes normality through a targeted modeling approach. This approach is based on explicitly phrased assumptions about how epileptogenic lesions are manifested in brain MRIs in contrast to non-lesional MRIs. These modeling assumptions are then used to guide the inductive bias of the deep learning architecture to identify deviations from those variances that are considered normal.

Informally speaking, abnormal, i.e. lesional brain MRIs, are distinguished from non-lesional MRIs by regions that involve only a small fraction of the overall brain volume. This assumption calls for presupposing a *local* notion of abnormality. As a first step towards translating this into a formal deep learning approach, we explicitly postulate the following assumptions about how epileptogenic lesions are distinguished in contrast to normal brain MRIs:

(A1) There is an upper limit of the brain volume that an EL can typically cover.

(A2) ELs are spatially surrounded by *normal* brain regions, so that the ELs deviate from the morphology that is to be expected from surrounding brain regions.

(A3) The MRI morphology of specific localized brain regions in *normal* MRIs is predictable from surrounding brain regions.

(A4) Brain MRIs can be registered into a coordinate system where the same brain regions across different brains are localized at similar coordinates.

The need for contextualizing normality can also be viewed from the perspective of machine learning theory. In supervised learning, it is a well-established consequence of the no-free-lunch theorem (Wolpert, 1996) that any learning algorithm must inevitably possess an inductive bias. As a consequence, (Sterkenburg and Grünwald, 2021) have raised the question of how to justify inductive bias. While these fundamentals of supervised machine learning have not been formally transferred to anomaly detection, related aspects such as notions of PAC-learnability and the role of the VC-dimension in anomaly detection have been investigated by Siddiqui et al. (2016). In this sense, the inductive bias of an anomaly detection implicitly contextualizes normality. For anomaly detection approaches without an explicitly contextualized normality, their inductive bias constitutes an implicit sense of normality, and contextualizing normality is one way to make anomaly detection more transparent.

In the SpyCAT approach proposed here, we justify inductive bias by the domain knowledge manifested in assumptions (A1)–(A4). This approach is also in line with recent work in supervised medical image analysis that adjusts inductive bias towards making deep learning models or their interpretable output compliant with existing domain knowledge (Maier et al., 2019; Mahapatra et al., 2022; Wang et al., 2024a; Schuhmacher et al., 2022).

## 4. Method

The SpyCAT approach translates assumptions (A1)–(A4) from Section 3 into an anomaly detection approach. Specifically, our method works by decomposing the complete 3D volume image ℐof a brain MRI into cubic patches. For a voxel position *p* within an image ℐ, we denote the patch originating at *p* by **I**_*p*_ ∈ ℝ ^*q*×*q*×*q*^, where throughout this manuscript we work with a patch size of *q* = 16, which matches assumption (A1). Our approach builds on inferring an anomaly map for individual patches, so that an anomaly map **A**_*map*_ for a complete MRI image ℐ can be assembled from a patch-decomposition.

In general, the basic idea is that a vector-quantized variational autoencoder (VQ-VAE) (Oord et al., 2017), which consists of an encoder *V*_enc_ : ℝ ^*q*×*q*×*q*^ ***→* E** and a corresponding decoder *V*_dec_, is trained to reliably compress and reconstruct either lesional or non-lesional input images **I**_*p*_ through a suitable embedding space **E**. Correspondingly, we expect to obtain an accurate reconstruction of **I**_*p*_ by computing 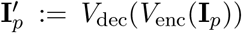. Following assumption (A2), we can set 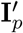 in contrast with a *counterfactual* version of the patch at position *p*, denoted by **C**_*p*_, that is to expected from neighboring brain regions, according to (A3). To obtain this counterfactual **C**_*p*_, we denote *n* as one patch position that is adjacent to position *p*, so that **I**_*n*_ denotes an image cube adjacent to **I**_*p*_. We then train a transformer network *T* that utilizes the encoded embedding of **I**_*n*_ as context information to predict the counterfactual embedding, so that

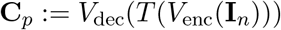

yields the desired counterfactual image that is expected from neighboring brain regions, as postulated in (A2). The difference image 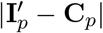 then yields the anomaly map at position *p*: If the patch at position *p* looks as expected from neighboring position *n* in a non-lesional brain MRI, then 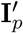 and **C**_*p*_ will coincide, yielding an all-zero anomaly map. If, however, a lesion is present at position *p*, the lesion will be missing in **C**_*p*_, thus uncovering the lesion in the difference image of the anomaly map.

As stated so far, the transformer *T* does not account for the brain region in which the embedding of the counterfactual image is to be generated. When dealing with brain MRIs registered into the coordinate system of a reference brain, the location information *p* is highly informative towards the morphology that is to be reconstructed, as postulated in assumption (A4). In a transformer network, this can be realized easily: We can simply pass the spatial patch coordinates *p* = (*x*_*p*_, *y*_*p*_, *z*_*p*_) as additional context information to the transformer, so that we obtain an improved counterfactual image from

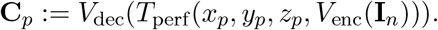

We additionally replaced the transformer architecture by an autoregressive performer (Choromanski et al., 2020), and denote this position-enhanced performer as *T*_perf_.

Overall, the training process for our complete anomaly detection network involves two steps. First, the VQ-VAE is trained to bring images into a compressed representation using an encoder, and also to obtain a decoder for reconstructing. In the second step, a transformer that learns lesion-free patches look like at a certain position in the image and corrects the generated embedding space accordingly. We have so far not yet discussed the embedding space **E** of the variational autoencoder. While the transformer (or performer) network could in principle operate on the level of raw or naively discretized image patches rather than in embedding space, the compression by a VAE has an important role in practice, to make the transformer network *T*_perf_ more data-efficient. As dicussed in the next paragraph, this requires an appropriate balance between a sufficient level of compression and a high reconstruction quality.

### Obtaining compressed patch representations

The key idea behind employing the VQ-VAE (Oord et al., 2017) is to learn a compressed, dictionary-like representation of patches in an embedding space **E** that facilitates accurate reconstruction of patches, which at the same time provides favorable context for a transformer network to predict the embedding of the neighboring patch. To this end, our main building blocks are *K* = 8192 codebook vectors *e*_0_, …, *e*_*K*−1_ ⊂ ℝ*^D^* with a feature depth of *D* = 256 to be inferred from training a VQ-VAE. The original VQ-VAE (Oord et al., 2017) represents the complete input image through a ℝ^*K*×*D*^ feature matrix, and maps an input to a single *D*-dimensional codebook vector through a one-hot-encoding of the rows of the feature matrix. In our setting, representing one input patch by a single codebook vector would be too coarse grained for the dedicated purpose of the representation, namely to predict and fully reconstruct a complete neighboring patch. As a more fine-grained and more informative representation, we train the encoder *V*_enc_ to yield a representation comprising 4 × 4 × 4 codebook vectors, along with a corresponding decoder *V*_dec_, which learns to reconstruct an image 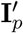 from the embedding space.

#### Algorithm 1

Generate Perlin Noise for Image Patch **I**_*p*_

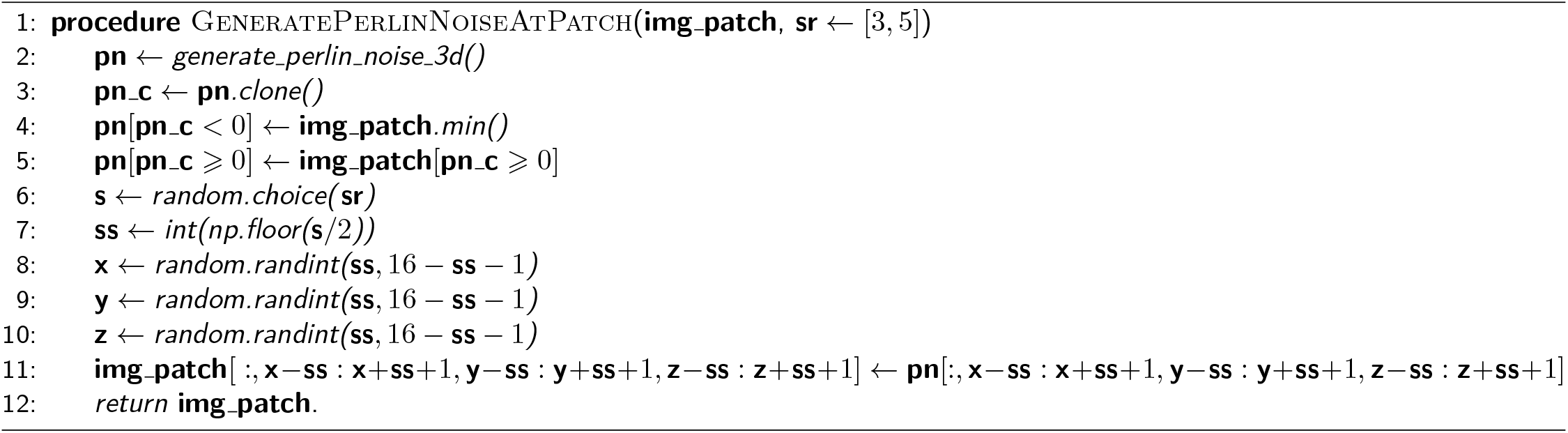

If we consider the codebook vectors *e*_0_, … *e*_*K*−1_ as fixed after training, we can now formally identify our compressed patch embedding space as

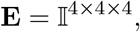

where I = {0, …, 8191] denotes the index space of the codebook vectors. In particular, this embedding space allows us to efficiently represent a single patch by 64 integer values in a way that, given the codebook vectors, the input patch can still be reconstructed accurately. At the same time, the representation based on codebook indices resembles token representations of large language models (Devlin et al., 2018; Touvron et al., 2023) and thus promises to work favorably as an input encoding for transformer networks.

### Generating pseudo-lesional patches

One issue to be addressed when training the autoencoder is that, for our approach to work as intended, *V*_enc_ and *V*_dec_ must be able to embed and reconstruct lesions. However, there is only a very small set of lesional patches available in the dataset, which is far from sufficient to learn their proper reconstruction. Furthermore, MRIs are very heterogeneous and they can have many artifacts. For example, in the SWI sequence, certain sections are commonly masked out (Schönlau, 2018). In order for such artifacts to be considered normal, they must also be present in the embedding space. In order to enhance the reconstruction capabilities beyond non-lesional patches, we augmented the data set by generating *pseudo-lesional* patches. To do so, we added random noise in the form of Perlin noise (Perlin, 1985) to each image patch **I**_*p*_. This allows us to use only MRIs marked as non-lesion MRIs for training the VQ-VAE and still achieve sufficient reconstructability at the patch level. In VQ-VAE training, image patches **I**_*p*_ are sampled at randomly chosen positions in the MRI image.

Perlin noise is a type of gradient noise that creates smooth transitions between points in the grid. For each patch, the function *GeneratePerlinNoiseAtPatch*() is called with **I**_*p*_ (see Figure 2a and Algorithm 1). Then 3D perlin noise is generated using the implementation of (Vigier, 2020) (see Figure 2b). All values greater than or equal to 0 are replaced with the image information from **I**_*p*_ and all values less than 0 are replaced with the minimum patch value **I**_*p*_ (see Figure 2c). A 3D box with side lengths of 3 or 5 is cut out at a random position in the resulting patch (from Figure 2c) and inserted at the same position in **I**_*p*_ (see Figure 2d).

**Figure 2:**
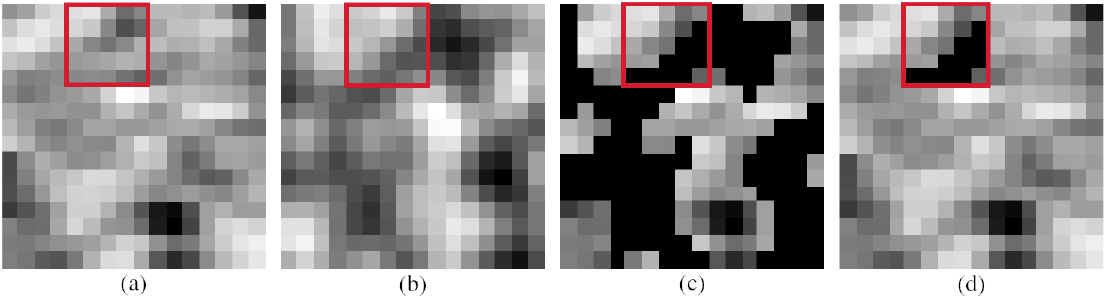
From left to right: (a) Image patch **I**_*p*_ with coronal view; (b) Perlin noise **pn**; (c) All values greater than or equal to 0 are replaced with the image information from **I**_*p*_ and all values less than 0 are replaced with the minimum patch value **I**_*p*_; (d) A 3D box with side lengths 3 or 5 (randomly chosen) is cut out of the image 2c and inserted into the original image **I**_*p*_ at the same position.

This creates small artifacts that resemble lesions at the patch level.

### Predicting counterfactual neighboring patches

The purpose of the VQ-VAE is to obtain a compressed patch representation which facilitates the transformer-based prediction of counterfactually non-lesional neighboring patches following assumptions (A2) and (A3).

We here employ the FAVOR+ algorithm introduced in the Performer Framework (Choromanski et al., 2020), which evades the quadratic complexity of the dependencies between query, key, and value vectors, and realizes an attention mechanism with linear time complexity. We train the transformer network *T*_perf_ on sequences composed of three parts, assuming that the latent representation of patch position *p* is to be predicted from a neighboring patch position *n*:

- The index coordinates *x*_*p*_, *y*_*p*_, *z*_*p*_ of the patch position *p* within the registered coordinate system of the underlying MRI image ℐ, which is encoded through a simple vocabulary embedding,
- the flattened latent space representation of the neighboring patch *z*_*n*_ := *V*_enc_(**I**_*n*_), and
- the flattened latent space representation of the patch *p* itself, i.e., *z*_*p*_ := *V*_enc_(**I**_*p*_).

Thus, each patch is represented by a sequence composed of 131 = 3+64+64 many tokens. During inference, only the position embedding and the neighborhood patch embedding are given, so the embedding *z*_*p*_ is predicted autoregressively, resulting in the counterfactual embedding.

The data for training *T*_perf_ are obtained by drawing image patches **I**_*p*_ at random positions *x*_*p*_, *y*_*p*_, *z*_*p*_ from the training dataset along with an image patch **I**_*n*_ at a neighboring position *p*. The patch size of 16 16 16 as well as the latent space dimension of 4 × 4 × 4 and the codebook size of *K* = 8192 were used constantly throughout all experiments.

To analyze a complete MRI image and determine whether a lesion is present or not, the image is decomposed into patches along a 16 × 16 × 16 voxel grid, where each point *p* on the grid yields an image patch **I**_*p*_ along with one neighboring patch **I**_*n*_. Each **I**_*p*_ and **I**_*n*_ is passed through *V*_enc_ to obtain embeddings **z**_*p*_ and **z**_*n*_, respectively. We then use *T*_perf_ to obtain a counterfactual embedding **z**_*c*_. The performer autoregressively replaces all elements given by *p* and **z**_*n*_ in the sequence that are below a threshold ***ϕ***. The resulting counterfactual embedding space **z**_*c*_ is then passed as input to *V*_dec_ along with the original embedding space **z**_*p*_. This produces two reconstructions 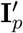 and **C**_*c*_, which are then simply subtracted from each other to obtain a patch anomaly map. All patch anomaly maps can be mapped back to their respective grid positions, which yields the whole-brain anomaly map **A**_map_.

## 5. Experiments

We evaluated SpyCAT on two dataset pairs: First, an in-house dataset of brain MRIs related to two types of epileptogenic lesions, henceforth referred to as the *AIM*.*E dataset*, and secondly, a dataset obtained by merging two public datasets with brain tumor related MRIs, henceforth referred to as the *IXI-BraTS dataset*. For each of the two datasets, we train one dedicated model. All non-lesion studies from the AIM.E dataset are used and evaluated on the lesion studies. The same applies to the IXI dataset.

### 5.1 Datasets

#### AIM.E dataset

The AIM.E dataset consists of MRIs that were collected in three hospitals (Bochum, Bottrop, Iserlohn) on different Siemens MRI scanners over a period of 11 years in the time peroid between 2010 and 2021. Each study contains at least one of the following sequences: 3D-T1 (1 × 1 × 1 mm), coronal and axial T2 (2 mm), 3D-Flair (1 × 1 × 1 mm) and axial SWI (2 mm) sequence (Wellmer et al., 2013). All MRI studies were evaluated by a physician by first categorizing them into non-lesional vs. lesional. Subsequently, lesions observed in the lesional MRIs were annotated with a singular reference point and given a class label identifying the type of lesion. Within the validation and test datasets, we only include studies where precisely one unambiguously labeled lesion is identified.

For training, only MRIs that have not been labeled *cavernoma* or *post resection cavity* are used. We consider these as *normal*. For validation and testing, we use lesional and *normal* MRIs. The exact division between training, validation and test dataset is shown in Figure 3. We consider the SWI sequence for cavernoma detection and the Flair sequence for post resection cavity detection (Wellmer et al., 2013; Wang et al., 2017). The division into training, validation, and test dataset takes place at study level, so that there are different numbers for Flair and SWI sequences.

**Figure 3:**
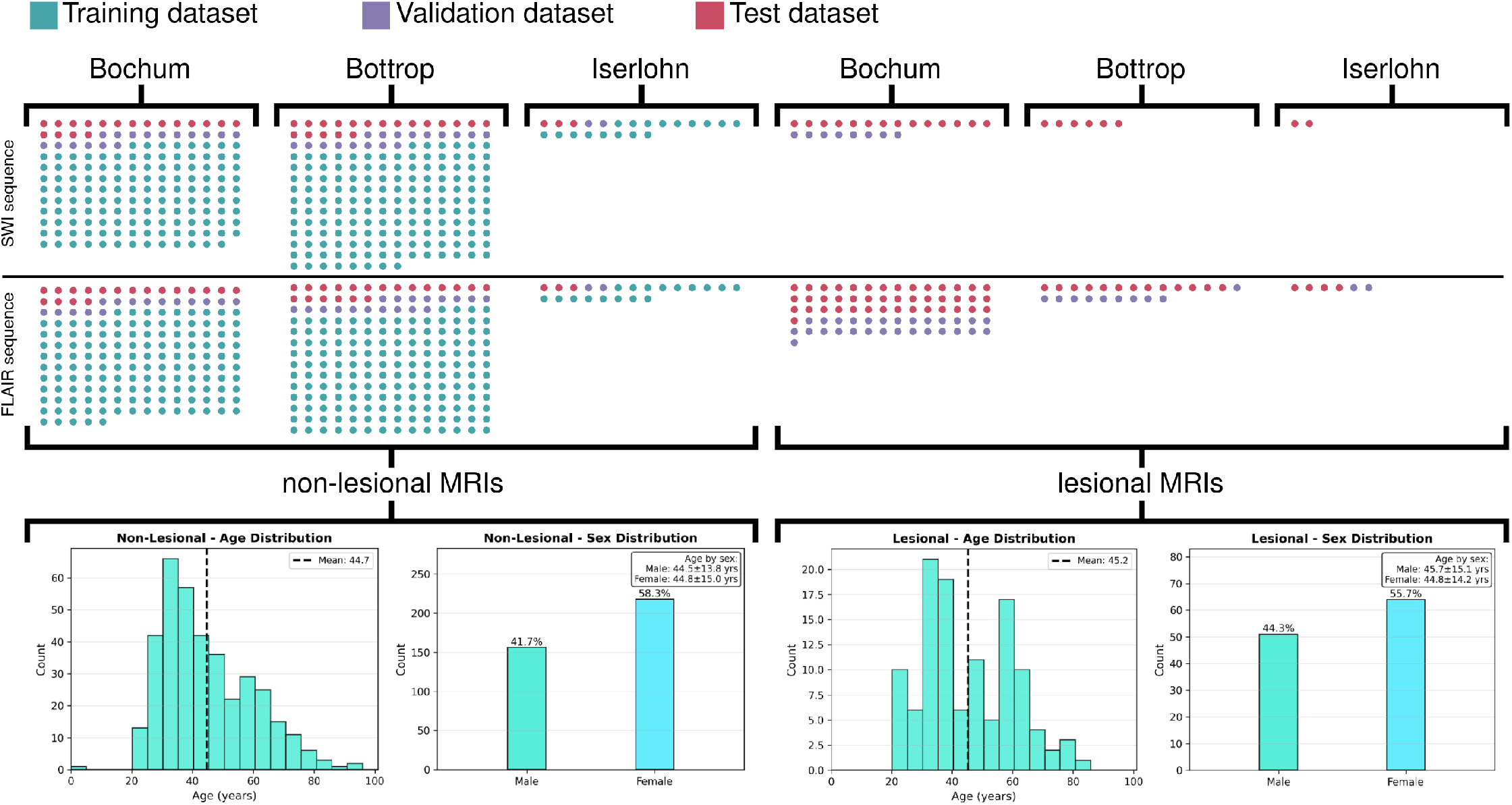
Division of the dataset into training, validation and test data. The MRIs are separated at study level and come from three different hospitals. SWI sequences are used for cavernoma detection and Flair sequences for post resection cavity detection.

#### IXI-BraTS-Dataset

We used the IXI dataset^2^ as a resource for obtaining 581 non-lesional MRI studies, each of contains a T1 and a T2 sequence. As a second resource, we utilized the BraTS dataset (Menze et al., 2015; Bakas et al., 2017, 2018), where each study consists of a T1, T2, and Flair sequence accompanied by a segmentation map for of five regions (no lesion, non-enhancing tumor core, peritumoral edema, and GD-enhancing tumor). All values greater than 0 are considered a lesion, which leads to binarization (0 non-lesional; 1 lesional). The lesion-free MRIs of the IXI dataset were used to train the VQ-VAE and the performer network, and the BraTS data was also used for evaluation. The IXI data were split into 60% training data, 20% validation data and 20% test data. The BraTS dataset was split into 50% each for validation and testing, respectively.

### 5.2 Preprocessing

The preprocessing pipeline consists of 4 steps: First, MRIs are registered on the MNI-152 template (Mandal et al., 2012) with a size of 193 × 229 × 193 using the SimpleITK frameworks versor transformation (Beare et al., 2018; Yaniv et al., 2018; Lowekamp et al., 2013). The T1 sequence of each study is taken and registered to the T1 sequence of the MRI template. The transformation matrix is saved and then applied to all other sequences in the study. Second, skull stripping is performed using the MNI-152 template mask. Third, we apply bias field correction using (Sled et al., 1998; Tustison et al., 2010). As a final pre-processing step, we perform histogram normalization following (Nyul et al., 2000).

### 5.3 Implementation details

#### Autoencoder

The topology of the autoencoder follows Oord et al. (2017) and consists of two convolutional layers with *LeakyReLU* activation function in the encoder *V*_enc_ and two transposed convolutional layers using *LeakyReLU* as activation function in the decoder *V*_dec_, each with dimensions 128 and 256 and six residual layers. We used the Adam optimizer (Kingma and Ba, 2014) for training with a learning rate of *l*_*V*_ = 10^−5^, a batch size of 256 and an exponential weight decay of *γ*_*V*_ = 0.95. Training is performed over 50 epochs, extracting 10, 000, 000 image patches during each epoch.

#### Transformer

The performer architecture in this paper is based on (Choromanski et al., 2020) and uses six layers, each with eight attention heads and 512 dimensions. The Adam optimizer (Kingma and Ba, 2014) is used with a learning rate of *l*_*T*_ = 10^−5^. The training is carried out over 50 epochs, during each of which 1, 000, 000 image patches **I**_*p*_ and neighborhood patches **I**_*n*_ are extracted. The batch size is 256. During the testing phase, the weights from the performer epoch that correspond to the highest validation AUROC metric were utilized.

### 5.4 Validation Approach

Validation involves the two main constituents of (i) identifying appropriate validation metrics and (ii) identifying appropriate reference methods.

#### Validation metrics

We used the *metrics reloaded* framework (Maier-Hein et al., 2024) as our basis for identifying appropriate validation measures. Since SpyCAT is an anonmaly detection approach, and due to the nature of epileptogenic lesions as well as the point-of-interest annotations available for the AIM.E dataset, none of the use cases from (Maier-Hein et al., 2024) are immediately applicable in our setting. Following the framework, we identified two relevant validation categories:

- *Image-level Classification (ImLC):* We measure ImLC performance of distinguishing non-lesional MRIs from lesional ones. We identified the Area Under Receiver Operating Curve (AUROC) as a suitable multi-threshold metric, using the mean across all voxels of the anomaly map **A**_map_ as a natural threshold variable, which we denote as *µ*_map_. Analogously to AUROC, we also report average precision (AP). Complementary to these two multi-threshold metrics, we also report accuracy (ACC), sensitivity (SENS) and specificity (SPEC) as single-threshold metrics. To obtain a single threshold, we determine the optimal threshold for *µ*_map_ by maximizing Youden’s J statistic, i.e., the difference between true positice rate and false positive rate, across the validation dataset. This threshold is then applied as a constant to the complete test dataset to calculate ACC (accuracy), SENS (sensitivity), and SPEC (specificity).
- *Object Detection (ObD):* Following the process from (Maier-Hein et al., 2024), we first define a *localization criterion*. We use different localization criteria for the BraTS dataset and the AIM.E dataset due to the very different nature of the lesions and the given annotations.

For the BraTS dataset, we note that well-established measures exist for validating supervised segmentation methods, which however are not applicable to the anomaly detection approaches investigated here. As a localization criterion for the BraTS dataset, we considered a tumor lesion as detected if any of the **k** highest intesity voxels of **A**_map_ is located within the ground truth segmentation. Different small values of **k** were tested through ablation, see Section 6.1 below.

For the AIM.E dataset, which lacks ground truth segmentations and possesses only point-of-interest annotations, an additional detection radius parameter needs to be introduced to obtain a localization criterion. Here, we consider a lesion to be detected if one of the **k** highest intesity voxels of **A**_map_ is located within a fixed radius of **r**. As for **k**, appropriate values for **r** are subject to ablation, see Section 6.1, where we varied **r** between **r** = 3 mm and **r** = 9 mm. This detection threshold radius is a much more conservative choice than in the related study by Spitzer et al. (2022), which tolerated a radius of **r** = 20 mm for the detection of focal cortical dysplasia lesions. We note that neither true negatives nor false positives can occur when evaluation *ObD*.

#### Reference methods

We employed two semi-supervised approaches as reference methods for comparing our approach to state-of-the-art: First, a 3D f-AnoGAN (Simarro et al., 2020), and second, a *β* -VAE (Loizillon et al., 2024). Both methods work at the whole-image level, which means that no patches are extracted, but images are processed as a whole. Both methods are also trained on non-lesional MRIs and tested on lesional MRIs (semi supervised approach). Both f-AnoGAN and *β* -VAE have the same underlying idea as our approach, namely that the lesional areas of the MRI, unlike *normal* healthy areas, cannot be reconstructed by the network, so an anomaly map is obtained by comparing the input image with the output reconstruction.

The implementation of the 3D f-AnoGAN utilized here is based on (Simarro et al., 2020), but does not reduce the original images to 64 × 64 × 64, but to 128 × 128 × 128, in order to have a chance to detect small lesions at all. In the implementation, a batch size of 2 is used. With the *β*-VAE, the full image resolution is used as input in contrast to the 3D f-AnoGAN. All other parameters follow (Simarro et al., 2020) and (Loizillon et al., 2024).

## 6. Results

To evaluate performance on the grounds of the *Metrics Reloaded* framework (Maier-Hein et al., 2024) on the two levels of *image classification* and *object detection*.

For the *image classification* case, Table 1 shows the number of true positives (TP) and false negatives (FN) for identifying cavernoma. In the lack of ground truth segmentations available for the AIM.E data set, we do not report a segmentation metric.

**Table 1:** Comparison of our approach with f-AnoGAN and *β*-VAE on the two pairs of datasets (IXI-BraTS and AIM.E dataset). On the IXI-BraTS dataset, MRIs of the T2 sequence are used. In the AIM.E dataset, the cavernoma evaluation (here abbreviated as *Cav*.) is performed on the SWI data and the post resection cavity evaluation (here abbreviated as *PRC*) is shown on the Flair sequence.

| Pair | Seq. | Model | Classification (*ImLC*) [%] |  |  |  |  | Detection (*ObD*) |  |  |
| --- | --- | --- | --- | --- | --- | --- | --- | --- | --- | --- |
|  |  |  | ACC | AUROC | AP | SENS | SPEC | TP | FN | TP + FN |
| IXI-BRATS-METS | T2 | f-AnoGAN | 74.69<sub>[74.8,79.3]</sub> | 81.50<sub>[81.6,84.6]</sub> | 85.94<sub>[86.0,88.1]</sub> | 65.08<sub>[65.5,73.3]</sub> | **87.42**<sub>[82.5,87.4]</sub> | 97 | 135 | 232 |
| | | β-VAE | 68.55<sub>[67.9,70.0]</sub> | 75.36<sub>[75.1,76.9]</sub> | 81.83<sub>[81.9,83.1]</sub> | 65.94<sub>[52.8,67.2]</sub> | 72.00<sub>[72.1,87.8]</sub> | 113 | 119 | 232 |
|  |  | DenosingAE | 57.27<sub>[55.3,60.1]</sub> | 34.65<sub>[34.7,37.9]</sub> | 47.73<sub>[46.3,47.8]</sub> | **99.14**<sub>[96.3,99.5]</sub> | 1.71<sub>[1.2,5.5]</sub> | 81 | 151 | 232 |
|  |  | PatchCore | 71.74<sub>[71.9,74.2]</sub> | 74.57<sub>[74.2,74.6]</sub> | 72.49<sub>[72.2,72.9]</sub> | 75.86<sub>[76.8,94.4]</sub> | 66.66<sub>[47.4,65.3]</sub> | 141 | 91 | 232 |
|  |  | **SpyCAT (Ours)** | **77.39**<sub>[73.2,77.9]</sub> | **85.22**<sub>[81.5,85.2]</sub> | **89.78**<sub>[86.9,89.8]</sub> | 72.41<sub>[65.0,73.2]</sub> | 83.99<sub>[78.3,84.9]</sub> | 168 | 64 | 232 |
| IXI-BraTS | T2 | f-AnoGAN | 71.25<sub>[70.8,75.5]</sub> | 76.55<sub>[75.6,82.6]</sub> | 84.18<sub>[83.4,87.7]</sub> | 61.21<sub>[61.2,66.9]</sub> | 84.57<sub>[83.5,86.7]</sub> | 152 | 80 | 232 |
| | | β-VAE | **80.83**<sub>[78.7,82.2]</sub> | **86.00**<sub>[85.0,87.4]</sub> | 91.11<sub>[90.7,92.0]</sub> | 72.48<sub>[68.9,75.6]</sub> | 91.14<sub>[86.1,92.0]</sub> | 197 | 35 | 232 |
|  |  | DenosingAE | 55.77<sub>[55.1,57.9]</sub> | 45.30<sub>[44.5,47.0]</sub> | 53.69<sub>[52.6,55.5]</sub> | **83.18**<sub>[74.6,89.7]</sub> | 19.42<sub>[15.6,29.2]</sub> | 131 | 101 | 232 |
|  |  | PatchCore | 70.27<sub>[70.4,74.3]</sub> | 75.88<sub>[75.7,78.2]</sub> | 77.31<sub>[76.6,79.6]</sub> | 68.10<sub>[68.3,93.3]</sub> | 73.14<sub>[49.3,73.1]</sub> | 191 | 41 | 232 |
|  |  | **SpyCAT (Ours)** | 78.38<sub>[78.8,89.5]</sub> | 85.74<sub>[86.0,94.4]</sub> | **91.14**<sub>[91.3,96.5]</sub> | 68.53<sub>[69.2,86.8]</sub> | **91.43**<sub>[91.5,93.1]</sub> | 210 | 22 | 232 |
| AIM.E-PRC | FLAIR | f-AnoGAN | 70.59<sub>[66.8,70.5]</sub> | 75.79<sub>[72.8,75.7]</sub> | 85.36<sub>[83.3,85.3]</sub> | 51.67<sub>[46.9,51.7]</sub> | **97.61**<sub>[95.2,97.5]</sub> | 25 | 35 | 60 |
| | | β-VAE | 69.60<sub>[64.9,69.6]</sub> | 75.91<sub>[73.1,75.8]</sub> | 83.36<sub>[79.5,83.2]</sub> | 58.33<sub>[47.3,58.3]</sub> | 85.71<sub>[83.5,90.2]</sub> | 37 | 23 | 60 |
|  |  | DenosingAE | 68.62<sub>[66.8,76.1]</sub> | 68.53<sub>[65.6,75.7]</sub> | 75.89<sub>[75.2,81.5]</sub> | 71.66<sub>[68.5,76.4]</sub> | 64.28<sub>[64.3,75.6]</sub> | 33 | 27 | 60 |
|  |  | PatchCore | 74.51<sub>[66.8,74.2]</sub> | **79.68**<sub>[71.8,79.4]</sub> | 83.82<sub>[78.5,83.6]</sub> | 70.00<sub>[53.3,71.2]</sub> | 79.10<sub>[77.2,81.2]</sub> | 43 | 17 | 60 |
|  |  | **SpyCAT (Ours)** | **75.49**<sub>[75.6,80.3]</sub> | 78.73<sub>[78.9,84.1]</sub> | **86.43**<sub>[84.3,86.7]</sub> | **71.67**<sub>[72.2,86.4]</sub> | 80.95<sub>[73.9,80.7]</sub> | 44 | 16 | 60 |
| AIM.E-Cav. | SWI | f-AnoGAN | 66.12<sub>[67.0,74.1]</sub> | 74.50<sub>[75.4,82.4]</sub> | 67.49<sub>[67.6,70.7]</sub> | **95.45**<sub>[90.9,95.2]</sub> | 0.50<sub>[50.7,65.0]</sub> | 18 | 4 | 22 |
| | | β-VAE | 70.97<sub>[71.1,77.3]</sub> | 83.00<sub>[83.5,83.9]</sub> | 74.66<sub>[74.7,76.1]</sub> | **95.45**<sub>[82.2,95.1]</sub> | 57.50<sub>[57.9,70.6]</sub> | 19 | 3 | 22 |
|  |  | DenosingAE | 74.19<sub>[72.7,74.2]</sub> | 62.73<sub>[62.8,65.5]</sub> | 56.46<sub>[49.7,56.3]</sub> | 50.00<sub>[50.2,54.5]</sub> | 87.50<sub>[82.6,87.4]</sub> | 16 | 6 | 22 |
|  |  | PatchCore | 77.42<sub>[77.6,82.2]</sub> | 73.63<sub>[73.9,83.2]</sub> | 59.36<sub>[59.6,72.7]</sub> | 68.18<sub>[68.6,81.6]</sub> | 82.49<sub>[80.1,84.9]</sub> | 18 | 4 | 22 |
|  |  | **SpyCAT (Ours)** | **83.87**<sub>[79.2,83.7]</sub> | **83.30**<sub>[79.0,83.2]</sub> | **79.39**<sub>[75.8,79.3]</sub> | 68.18<sub>[68.4,72.7]</sub> | **92.50**<sub>[82.7,92.3]</sub> | 21 | 1 | 22 |

Figure 4 shows the three classes to be examined, namely *cavernomas, post resection cavity defects* as well as *tumor* in the BraTS data. For each entity, an example image ℐ (coronal view), an anomaly map **A**_map_ of the network, which is composed of the individual image patches, and a ground truth map with a point-of-interest annotation (AIM.E dataset) or a ground truth segmentation map (BraTS dataset) are shown.

**Figure 4:**
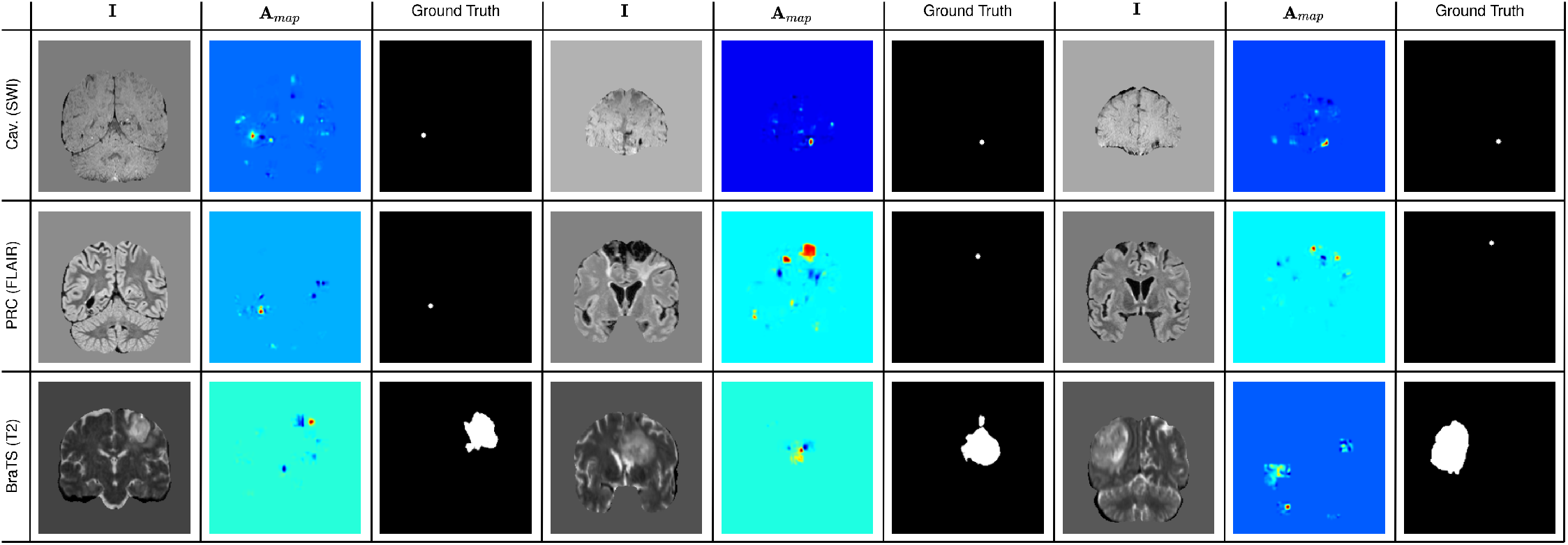
Representation of the output of the presented model on the three identities cavernoma, post resection cavity and tumor data. Ground truth point-of-interest annotations exist for the AIM.E data and a segmentation map exists for the BraTS dataset. For all input images ℐ the output anomaly map **A**_*map*_, which is composed of the individual patches, are shown.

It is easy to see that even the smallest lesions (cavernomas and post resection cavity) are highlighted well in the anomaly map **A**_*map*_. Here it is particularly clear that a deviation is visible in contrast to the local neighborhood patch **I**_*n*_ and the position embedding *x*_*p*_, *y*_*p*_, *z*_*p*_. For large-area lesions, such as those found in the BraTS tumor dataset, tumor tissue is also present in the neighboring patch **I**_*n*_, and the performer subsequently reconstructs tumor tissue as well. The patch position *x*_*p*_, *y*_*p*_, *z*_*p*_ also does not seem to provide suffient context to detect the anomaly. Despite these, tumors are also detected in the BraTS dataset. This can be specifically attributed to the fact that brain tumors do not fully match assumptions (A1), (A2) and (A3), to which our approach has been tailored.

### 6.1 Ablation Studies

All ablation studies were performed on the cavernoma data using the SWI sequence.

#### Detection threshold *ϕ*

It is particularly important to determine when the performer considers an element in the sequence to be abnormal, given the position embedding *x*_*p*_, *y*_*p*_, *z*_*p*_ and the neighborhood sequence **z**_*n*_. Since there is no straightforward choice of ***ϕ*** that can be justified mathematically or by domain knowledge, we treat ***ϕ*** as a parameter. The parameter ***ϕ*** determines which elements in the sequence are replaced and which are not. If the value of the element falls below ***ϕ***, then the element is replaced by the element suggested by the performer. In this ablation study, we tested three values ***ϕ*** ∈ {0.01, 0.001, 0.0001}. The results shown in Table 2 indicate that ***ϕ*** should be adjusted to specific image data sets.

**Table 2:** Results of the ablation studies for the parameters ***ϕ*** ∈ {0.01, 0.001, 0.0001}, **r** = {3, 5, 7, 9}, **k** = {3, 5, 10} and training of the VQ-VAE with (y) and without perlin (n) noise **pn**. Note that modifying **r** and **k** affects detection, but not classification, so that their ablation in the classification columns remains blank.

| Pair | Seq. | $\phi$ | r | k | pn | q | ovlp | Classification [%] | | | | | Detection | | |
| --- | --- | --- | --- | --- | --- | --- | --- | --- | --- | --- | --- | --- | --- | --- | --- |
|  |  |  |  |  |  |  |  | ACC | AUROC | AP | SENS | SPEC | TP | FN | TP + FN |
| AIM.E-Cav. | SWI | $\phi = 0.01$ | r = 9 | k = 10 | y | q = 16 | 0% | 72.58<sub>[68.0,75.6]</sub> | 76.02<sub>[73.6,81.2]</sub> | 71.25<sub>[67.6,76.5]</sub> | 72.73<sub>[73.0,85.9]</sub> | 72.50<sub>[62.9,72.4]</sub> | 17 | 5 | 22 |
| | | $\phi = 0.001$ | r = 9 | k = 10 | y | q = 16 | 0% | 83.87<sub>[79.2,83.7]</sub> | 83.30<sub>[79.0,83.2]</sub> | 79.39<sub>[75.8,79.3]</sub> | 68.18<sub>[68.4,72.7]</sub> | 92.50<sub>[82.7,92.3]</sub> | 21 | 1 | 22 |
| | | $\phi = 0.0001$ | r = 9 | k = 10 | y | q = 16 | 0% | 79.03<sub>[79.3,86.9]</sub> | 79.32<sub>[79.7,89.8]</sub> | 78.30<sub>[78.7,87.2]</sub> | 68.18<sub>[68.2,81.1]</sub> | 85.00<sub>[85.0,96.9]</sub> | 16 | 6 | 22 |
| | | $\phi = 0.001$ | r = 3 | k = 10 | y | q = 16 | 0% | | | | | | 15 | 7 | 22 |
| | | $\phi = 0.001$ | r = 5 | k = 10 | y | q = 16 | 0% | | | | | | 20 | 2 | 22 |
| | | $\phi = 0.001$ | r = 7 | k = 10 | y | q = 16 | 0% | | | | | | 20 | 2 | 22 |
| | | $\phi = 0.001$ | r = 9 | k = 10 | y | q = 16 | 0% | | | | | | 21 | 1 | 22 |
| | | $\phi = 0.001$ | r = 9 | k = 10 | y | q = 16 | 0% | | | | | | 21 | 1 | 22 |
| | | $\phi = 0.001$ | r = 9 | k = 5 | y | q = 16 | 0% | | | | | | 19 | 3 | 22 |
| | | $\phi = 0.001$ | r = 9 | k = 3 | y | q = 16 | 0% | | | | | | 18 | 4 | 22 |
| | | $\phi = 0.001$ | r = 9 | k = 10 | y | q = 16 | 0% | 83.87<sub>[79.2,83.7]</sub> | 83.30<sub>[79.0,83.2]</sub> | 79.39<sub>[75.8,79.3]</sub> | 68.18<sub>[68.4,72.7]</sub> | 92.50<sub>[82.7,92.3]</sub> | 21 | 1 | 22 |
| | | $\phi = 0.001$ | r = 9 | k = 10 | n | q = 16 | 0% | 79.03<sub>[71.2,78.8]</sub> | 76.59<sub>[72.1,76.5]</sub> | 74.28<sub>[68.2,74.1]</sub> | 59.09<sub>[57.2,61.0]</sub> | 90.00<sub>[86.2,91.9]</sub> | 19 | 3 | 22 |
| | | $\phi = 0.001$ | r = 9 | k = 10 | y | q = 16 | 0% | 83.87<sub>[79.2,83.7]</sub> | 83.30<sub>[79.0,83.2]</sub> | 79.39<sub>[75.8,79.3]</sub> | 68.18<sub>[68.4,72.7]</sub> | 92.50<sub>[82.7,92.3]</sub> | 21 | 1 | 22 |
| | | $\phi = 0.001$ | r = 9 | k = 10 | y | q = 12 | 0% | 74.19<sub>[72.7,83.4]</sub> | 76.93<sub>[77.1,80.5]</sub> | 70.47<sub>[70.7,79.4]</sub> | 77.27<sub>[65.0,81.6]</sub> | 72.50<sub>[67.5,90.6]</sub> | 20 | 2 | 22 |
| | | $\phi = 0.001$ | r = 9 | k = 10 | y | q = 8 | 0% | 61.29<sub>[59.8,61.5]</sub> | 67.72<sub>[65.8,67.8]</sub> | 53.99<sub>[52.8,57.7]</sub> | 90.90<sub>[90.9,95.2]</sub> | 45.00<sub>[40.3,45.0]</sub> | 17 | 5 | 22 |
| | | $\phi = 0.001$ | r = 9 | k = 10 | y | q = 16 | 0% | 83.87<sub>[79.2,83.7]</sub> | 83.30<sub>[79.0,83.2]</sub> | 79.39<sub>[75.8,79.3]</sub> | 68.18<sub>[68.4,72.7]</sub> | 92.50<sub>[82.7,92.3]</sub> | 21 | 1 | 22 |
| | | $\phi = 0.001$ | r = 9 | k = 10 | y | q = 16 | 25% | 85.48<sub>[81.6,85.3]</sub> | 84.63<sub>[81.2,84.5]</sub> | 82.61<sub>[77.7,82.5]</sub> | 69.09<sub>[68.3,71.4]</sub> | 94.12<sub>[87.7,94.8]</sub> | 21 | 1 | 22 |
| | | $\phi = 0.001$ | r = 9 | k = 10 | y | q = 16 | 50% | 87.09<sub>[84.0,87.0]</sub> | 85.97<sub>[83.4,85.7]</sub> | 85.83<sub>[79.6,85.7]</sub> | 70.01<sub>[68.2,70.1]</sub> | 95.75<sub>[92.6,97.4]</sub> | 21 | 1 | 22 |

#### Validation radius r

Another important parameter in our validation procedure is when a lesion is recognized as a lesion, i.e., which radius **r** is chosen. In the lack of a natural choice for **r**, we evaluated the influence of different radii **r** ∈ {3 mm, 5 mm, 7 mm, 9 mm}. The larger the lesion, the larger **r** must be chosen: Since only one point-of-interest annotation is available, the limit is not always easy to agree on. In general, we have followed the procedure of (Spitzer et al., 2023), where a radius of **r** = 20 mm is chosen for the detection of focal cortical dysplasia lesions.

### Validation top k criterion

The choice of the parameter **k**, which describes up to which number of the top *k* values of the anomaly map **A**_*map*_ a lesion is considered to be detected, is another validation parameter that does not follow a natural choice. We therefore tested the values **k** ∈ {3, 5, 10}, see Table 2.

#### Perlin-Noise

In the training process of the VQ-VAE, we use Perlin-Noise as data augmentation with the intuition that the autoencoder also learns with the data augmentation to restore *lesion-like* tissue and to map it in the embedding space. Comparison with and without the data augmentation can be seen in Table 2.

## 7. Discussion and Conclusion

With SpyCAT, we have presented a novel anomaly detection approach that identifies anomalies as deviations from local context, which is realized by combining a transformer-based approach with patch tokenization through a vector quantised variational autoencoder. We demonstrates that our approach improves the performance of conceptually related state-of-the-art approaches.

The detection of epileptogenic lesions has caught considerable attention in recent years. As reviewed by Walger et al. (2023), major efforts have been spent on the identification of focal cortical displasias (FCDs) and their multicentric validation (Kersting et al., 2024). While FCDs have not been included in our present study, the explicit assumptions underlying our anomaly detection apply to to FCDs as well, so that it is promising to assess FCD detection using our proposed approach.

It is generally conceivable to involve further spatial information by feeding symmetric context, in addition to or as replacement of neighbouring context, into the transformer model. In general, the brain is highly symmetric, so that symmetric context should be highly predictive for most brain regions. Attempts to realize this in practice, however, have failed in our current approach (data not shown). One likely reason is the insufficient precision of registration to the MNI brain, which may be worthwhile to assess under promising recent progress in brain registration (Wang et al., 2024b).

While SpyCAT detects lesions more reliably than reference methods within a suitable detection radius, it is in its current form not able to provide segmentations of the lesions, but only yields localization at the patch level. In principle, this could be addressed by striding the detection patch voxel by voxel, this is computationally prohibitively expensive in practice. For our purpose, namely the detection of small lesions, localization at the patch level can be considered sufficient and comes at the further advantage that for validation, only point annotations are required.

A methodological contribution of SpyCAT beyond brain MRI image analysis is the use of the VQ-VAE to produce discrete embeddings as encodings of 3D patches. Since it is plausible that transformer networks perform favorable on discrete tokens, this idea appears natural and has been described in similar form for the very different task of modeling of episodic memory in cognitive science (Fayyaz et al., 2022).

Our approach is based on explicit modeling assumptions about what kinds of variances set normal images apart from abnormal ones. In this respect, our approach deviates significantly from most previous anomaly detection methods, which tend to be conceived as generic and application independent approaches. Our approach builds on the assumption that any anomaly detection algorithm, just like any supervised learning algorithm, inevitably involves if not explicit, then implicit assumptions in the form of an inductive bias. While for supervised learning, this is well-established through the no-free-lunch theorems of supervised learning, these theoretical aspects have not been studied for anomaly detection. In this direction, our work calls for investigating in how far and in what form anomaly detection is subject to no-free-lunch theorems.

One practically relevant implication of our modeling-based approach is that the explicit assumptions facilitate explanation of misidentified anomalies. For example, we can explain why only small subregions of the relatively large tumors in the BraTS data set are identified by SpyCAT: major fractions of tumors cover several adjacent patches and thus are inconsistent with our prior assumptions. Similarly, it could in principle be possible to attribute other false negative identifications of patches to further deviations in the neighboring patches, which then fail to properly identify and localize the abnormality. In this sense, the explicit underlying assumptions of our approach warrant a certain degree of explainability, and thus improve the transparency compared to approaches without explixit assumptions.

## Data Availability

All data produced in the present study are available upon reasonable request to the authors

## Acknowledgments

This research was funded in part by the humAIne project, funded by the German Ministry of Science and Education (FKZ 02L19C203) and the FoRUM program of the Medical Faculty of the Ruhr University Bochum (F1089N-2004). We thank Sven Kreienbrock and Tobias Erm for technical assistance.

## Ethical Standards

The study was approved by the Ethics Committee of the medical faculty of Ruhr-University Bochum (21-7336).

## Data availability

The AIM.E dataset is available on request from the authors.

## Footnotes

1 https://brain-development.org/ixi-dataset/

2 https://brain-development.org/ixi-dataset/

## References

Spyridon Bakas, Hamed Akbari, Aristeidis Sotiras, Michel Bilello, Martin Rozycki, Justin S. Kirby, John B. Freymann, Keyvan Farahani, and Christos Davatzikos. Advancing the cancer genome atlas glioma mri collections with expert segmentation labels and radiomic features. Scientific data, 4:170117, September 2017. ISSN 2052-4463.

Spyridon Bakas, Mauricio Reyes, Andras Jakab, Stefan Bauer, Markus Rempfler, Alessandro Crimi, Russell Takeshi Shinohara, Christoph Berger, Sung Min Ha, Martin Rozycki, et al. Identifying the best machine learning algorithms for brain tumor segmentation, progression assessment, and overall survival prediction in the brats challenge. arXiv preprint arXiv:1811.02629, 2018.

Christoph Baur, Benedikt Wiestler, Mark Muehlau, Claus Zimmer, Nassir Navab, and Shadi Albarqouni. Modeling healthy anatomy with artificial intelligence for unsupervised anomaly detection in brain MRI. Radiology: Artificial Intelligence, 3(3):e190169, 2021.

Richard Beare, Bradley Lowekamp, and Ziv Yaniv. Image segmentation, registration and characterization in R with SimpleITK. Journal of Statistical Software, 86(8):1âĂŞ35, 2018. URL https://www.jstatsoft.org/index.php/jss/article/view/v086i08.

Finn Behrendt, Debayan Bhattacharya, Julia Krüger, Roland Opfer, and Alexander Schlaefer. Patched diffusion models for unsupervised anomaly detection in brain MRI. March 2023.

Paul Bergmann, Michael Fauser, David Sattlegger, and Carsten Steger. Mvtec ad–a comprehensive real-world dataset for unsupervised anomaly detection. In Proceedings of the IEEE/CVF conference on computer vision and pattern recognition, pages 9592–9600, 2019.

Simona Bottani, Ninon Burgos, Aurelien Maire, Dario Saracino, Sebastian Ströer, Didier Dormont, and Olivier Colliot. Evaluation of MRI-based machine learning approaches for computer-aided diagnosis of dementia in a clinical data warehouse. Medical Image Analysis, 89:102903, 2023. ISSN 1361-8415.. URL https://www.sciencedirect.com/science/article/pii/S1361841523001639.

Christopher Bowles, Chen Qin, Ricardo Guerrero, Roger Gunn, Alexander Hammers, David Alexander Dickie, Maria Valdés Hernández, Joanna Wardlaw, and Daniel Rueckert. Brain lesion segmentation through image synthesis and outlier detection. NeuroImage: Clinical, 16: 643–658, 2017.

Aaron Carass, Snehashis Roy, Amod Jog, Jennifer L Cuzzocreo, Elizabeth Magrath, Adrian Gherman, Julia Button, James Nguyen, Ferran Prados, Carole H Sudre, et al. Longitudinal multiple sclerosis lesion segmentation: resource and challenge. NeuroImage, 148:77–102, 2017.

Krzysztof Choromanski, Valerii Likhosherstov, David Dohan, Xingyou Song, Andreea Gane, Tamas Sarlos, Peter Hawkins, Jared Davis, Afroz Mohiuddin, Lukasz Kaiser, David Belanger, Lucy Colwell, and Adrian Weller. Rethinking attention with performers. September 2020.

Thomas Defard, Aleksandr Setkov, Angelique Loesch, and Romaric Audigier. PaDiM: a patch distribution modeling framework for anomaly detection and localization. November 2020.

Jacob Devlin, Ming-Wei Chang, Kenton Lee, and Kristina Toutanova. Bert: Pre-training of deep bidirectional transformers for language understanding. October 2018.

Nima Fathi, Amar Kumar, Brennan Nichyporuk, Mohammad Havaei, and Tal Arbel. Decodex: Confounder detector guidance for improved diffusion-based counterfactual explanations. arXiv preprint arXiv:2405.09288, 2024.

Gian Mario Favero, Parham Saremi, Emily Kaczmarek, Brennan Nichyporuk, and Tal Arbel. Conditional diffusion models are medical image classifiers that provide explainability and uncertainty for free. arXiv preprint arXiv:2502.03687, 2025.

Zahra Fayyaz, Aya Altamimi, Carina Zoellner, Nicole Klein, Oliver T Wolf, Sen Cheng, and Laurenz Wiskott. A model of semantic completion in generative episodic memory. Neural Computation, 34(9):1841–1870, 2022.

Ravnoor Singh Gill, Hyo-Min Lee, Benoit Caldairou, Seok-Jun Hong, Carmen Barba, Francesco Deleo, Ludovico D’Incerti, Vanessa Cristina Mendes Coelho, Matteo Lenge, Mira Semmelroch, Dewi Victoria Schrader, Fabrice Bartolomei, Maxime Guye, Andreas Schulze-Bonhage, Horst Urbach, Kyoo Ho Cho, Fernando Cendes, Renzo Guerrini, Graeme Jackson, R Edward Hogan, Neda Bernasconi, and Andrea Bernasconi. Multicenter validation of a deep learning detection algorithm for focal cortical dysplasia. Neurology, 97(16):e1571–e1582, October 2021.

Jia Guo, Shuai Lu, Lize Jia, Weihang Zhang, and Huiqi Li. Encoder-decoder contrast for unsupervised anomaly detection in medical images. IEEE Transactions on Medical Imaging, 43(3):1102–1112, 2024.

Ahmed Hamada. Br35h: Brain tumor detection 2020, 2020. URL https://www.kaggle.com/ahmedhamada0/brain-tumor-detection.

Hasan Iqbal, Umar Khalid, Jing Hua, and Chen Chen. Unsupervised anomaly detection in medical images using masked diffusion model. May 2023.

Antanas Kascenas, Rory Young, BjÃÿrn Sand Jensen, Nicolas Pugeault, and Alison Q. OâĂŹNeil. Anomaly detection via context and local feature matching. In 2022 IEEE 19th International Symposium on Biomedical Imaging (ISBI), pages 1–5, 2022.

Lennart N. Kersting, Lennart Walger, Tobias Bauer, Vadym Gnatkovsky, Fabiane Schuch, Bastian David, Elisabeth Neuhaus, Fee Keil, Anna Tietze, Felix Rosenow, Angela M. Kaindl, Elke Hattingen, Hans-Jürgen Huppertz, Alexander Radbruch, Rainer Surges, and Theodor Rüber. Detection of focal cortical dysplasia: Development and multicentric evaluation of artificial intelligence models. Epilepsia, n/a(n/a), 2024. URL https://onlinelibrary.wiley.com/doi/abs/10.1111/epi.18240.

Diederik P. Kingma and Jimmy Ba. Adam: A method for stochastic optimization. December 2014.

Sungwook Lee, Seunghyun Lee, and Byung Cheol Song. CFA: Coupled-hypersphere-based feature adaptation for target-oriented anomaly localization. June 2022.

Sophie Loizillon, Yannick Jacob, Maire Aurélien, Didier Dormont, Olivier Colliot, Ninon Burgos, and APPRIM-AGE Study Group. Detecting brain anomalies in clinical routine with the β-VAE: Feasibility study on age-related white matter hyperintensities. In Medical Imaging with Deep Learning, 2024.

Bradley C Lowekamp, David T Chen, Luis Ibáñez, and Daniel Blezek. The design of SimpleITK. Frontiers in neuroinformatics, 7:45, 2013.

Guoting Luo, Wei Xie, Ronghui Gao, Tao Zheng, Lei Chen, and Huaiqiang Sun. Unsupervised anomaly detection in brain mri: Learning abstract distribution from massive healthy brains. Computers in Biology and Medicine, 154:106610, 2023. ISSN 0010-4825. URL https://www.sciencedirect.com/science/article/pii/S0010482523000756.

Dwarikanath Mahapatra, Alexander Poellinger, and Mauricio Reyes. Interpretability-guided inductive bias for deep learning based medical image. Medical image analysis, 81:102551, 2022.

Andreas K Maier, Christopher Syben, Bernhard Stimpel, Tobias Würfl, Mathis Hoffmann, Frank Schebesch, Weilin Fu, Leonid Mill, Lasse Kling, and Silke Christiansen. Learning with known operators reduces maximum error bounds. Nature machine intelligence, 1(8):373–380, 2019.

Lena Maier-Hein, Annika Reinke, Patrick Godau, Minu D Tizabi, Florian Buettner, Evangelia Christodoulou, Ben Glocker, Fabian Isensee, Jens Kleesiek, Michal Kozubek, et al. Metrics reloaded: recommendations for image analysis validation. Nature methods, 21(2):195–212, 2024.

Pravat K. Mandal, Rashima Mahajan, and Ivo D. Dinov. Structural brain atlases: design, rationale, and applications in normal and pathological cohorts. Journal of Alzheimer’s disease : JAD, 31 Suppl 3:S169–S188, 2012. ISSN 1875-8908.

Sergio Naval Marimont and Giacomo Tarroni. Anomaly detection through latent space restoration using vector-quantized variational autoencoders. December 2020.

Felix Meissen, Johannes Paetzold, Georgios Kaissis, and Daniel Rueckert. Unsupervised anomaly localization with-ÂĂstructural feature-autoencoders. In Spyridon Bakas, Alessandro Crimi, Ujjwal Baid, Sylwia Malec, Monika Pytlarz, Bhakti Baheti, Maximilian Zenk, and Reuben Dorent, editors, Brainlesion: Glioma, Multiple Sclerosis, Stroke and Traumatic Brain Injuries, pages 14–24, Cham, 2023. Springer Nature Switzerland. ISBN 978-3-031-33842-7.

Bjoern H. Menze, Andras Jakab, Stefan Bauer, Jayashree Kalpathy-Cramer, Keyvan Farahani, Justin Kirby, Yuliya Burren, Nicole Porz, Johannes Slotboom, Roland Wiest, Levente Lanczi, Elizabeth Gerstner, Marc-AndrÃľ Weber, Tal Arbel, Brian B. Avants, Nicholas Ayache, Patricia Buendia, D. Louis Collins, Nicolas Cordier, Jason J. Corso, Antonio Criminisi, Tilak Das, HervÃľ Delingette, ÃĞaÄ§atay Demiralp, Christopher R. Durst, Michel Dojat, Senan Doyle, Joana Festa, Florence Forbes, Ezequiel Geremia, Ben Glocker, Polina Golland, Xiaotao Guo, Andac Hamamci, Khan M. Iftekharuddin, Raj Jena, Nigel M. John, Ender Konukoglu, Danial Lashkari, JosÃľ AntoniÃş Mariz, Raphael Meier, SÃľrgio Pereira, Doina Precup, Stephen J. Price, Tammy Riklin Raviv, Syed M. S. Reza, Michael Ryan, Duygu Sarikaya, Lawrence Schwartz, Hoo-Chang Shin, Jamie Shotton, Carlos A. Silva, Nuno Sousa, Nagesh K. Subbanna, Gabor Szekely, Thomas J. Taylor, Owen M. Thomas, Nicholas J. Tustison, Gozde Unal, Flor Vasseur, Max Wintermark, Dong Hye Ye, Liang Zhao, Binsheng Zhao, Darko Zikic, Marcel Prastawa, Mauricio Reyes, and Koen Van Leemput. The multimodal brain tumor image segmentation benchmark (BRATS). IEEE transactions on medical imaging, 34:1993–2024, October 2015. ISSN 1558-254X.

L.G. Nyul, J.K. Udupa, and Xuan Zhang. New variants of a method of mri scale standardization. IEEE Transactions on Medical Imaging, 19(2):143–150, 2000.

Aaron van den Oord, Oriol Vinyals, and Koray Kavukcuoglu. Neural discrete representation learning. November 2017.

Ken Perlin. An image synthesizer. ACM Siggraph Computer Graphics, 19(3):287–296, 1985.

Karsten Roth, Latha Pemula, Joaquin Zepeda, Bernhard SchÃűlkopf, Thomas Brox, and Peter Gehler. Towards total recall in industrial anomaly detection. June 2021.

Thomas Schlegl, Philipp Seeböck, Sebastian M. Waldstein, Georg Langs, and Ursula Schmidt-Erfurth. f-anogan: Fast unsupervised anomaly detection with generative adversarial networks. Medical Image Analysis, 54:30–44, 2019. ISSN 1361-8415. URL https://www.sciencedirect.com/science/article/pii/S1361841518302640.

Lars Schönlau. Optimale räumliche Orientierung und Angulierung von T2*-und SWI-Sequenzen im Rahmen eines epilepsiespezifischen MRT-Protokolls. doctoralthesis, Ruhr-Universität Bochum, Universitätsbibliothek, 2018.

David Schuhmacher, Stephanie Schörner, Claus Küpper, Frederik GroÃ§erueschkamp, Carlo Sternemann, Celine Lugnier, Anna-Lena Kraeft, Hendrik Jtte, Andrea Tannapfel, Anke Reinacher-Schick, Klaus Gerwert, and Axel Mosig. A framework for falsifiable explanations of machine learning models with an application in computational pathology. Medical image analysis, 82:102594, August 2022. ISSN 1361-8423.

Johannes Schwarz, Lena Will, Jörg Wellmer, and Axel Mosig. A patch-based student-teacher pyramid matching approach to anomaly detection in 3d magnetic resonance imaging. In Medical Imaging with Deep Learning, 2024. URL https://openreview.net/pdf?id=vh01Nd5PCl.

Md Amran Siddiqui, Alan Fern, Thomas G Dietterich, and Shubhomoy Das. Finite sample complexity of rare pattern anomaly detection. In UAI, volume 16, pages 686–695, 2016.

Jaime Simarro, Ezequiel de la Rosa, Thijs Vande Vyvere, David Robben, and Diana M. Sima. Unsupervised 3d brain anomaly detection. In: Brainlesion: Glioma, Multiple Sclerosis, Stroke and Traumatic Brain Injuries. BrainLes 2020. Lecture Notes in Computer Science, vol 12658. Springer, Cham (2021), October 2020.

J.G. Sled, A.P. Zijdenbos, and A.C. Evans. A nonparametric method for automatic correction of intensity nonuniformity in mri data. IEEE Transactions on Medical Imaging, 17(1):87–97, 1998.

Hannah Spitzer, Mathilde Ripart, Kirstie Whitaker, Felice D’Arco, Kshitij Mankad, Andrew A Chen, Antonio Napolitano, Luca De Palma, Alessandro De Benedictis, Stephen Foldes, Zachary Humphreys, Kai Zhang, Wenhan Hu, Jiajie Mo, Marcus Likeman, Shirin Davies, Christopher Güttler, Matteo Lenge, Nathan T Cohen, Yingying Tang, Shan Wang, Aswin Chari, Martin Tisdall, Nuria Bargallo, Estefanía Conde-Blanco, Jose Carlos Pariente, Saül Pascual-Diaz, Ignacio Delgado-Martínez, Carmen Pérez-Enríquez, Ilaria Lagorio, Eugenio Abela, Nandini Mullatti, Jonathan O’Muircheartaigh, Katy Vecchiato, Yawu Liu, Maria Eugenia Caligiuri, Ben Sinclair, Lucy Vivash, Anna Willard, Jothy Kandasamy, Ailsa McLellan, Drahoslav Sokol, Mira Semmelroch, Ane G Kloster, Giske Opheim, Letícia Ribeiro, Clarissa Yasuda, Camilla Rossi-Espagnet, Khalid Hamandi, Anna Tietze, Carmen Barba, Renzo Guerrini, William Davis Gaillard, Xiaozhen You, Irene Wang, Sofía González-Ortiz, Mariasavina Severino, Pasquale Striano, Domenico Tortora, Reetta Kälviäinen, Antonio Gambardella, Angelo Labate, Patricia Desmond, Elaine Lui, Terence O’Brien, Jay Shetty, Graeme Jackson, John S Duncan, Gavin P Winston, Lars H Pinborg, Fernando Cendes, Fabian J Theis, Russell T Shinohara, J Helen Cross, Torsten Baldeweg, Sophie Adler, and Konrad Wagstyl. Interpretable surface-based detection of focal cortical dysplasias: a multi-centre epilepsy lesion detection study. Brain, 145(11):3859–3871, November 2022.

Hannah Spitzer, Mathilde Ripart, Abdulah Fawaz, Logan Z. J. Williams, Emma C. Robinson, Juan Eugenio Iglesias, Sophie Adler, and Konrad Wagstyl. Robust andÂĂgeneralisable segmentation ofÂĂsubtle epilepsy-causing lesions: A graph convolutional approach. In Hayit Greenspan, Anant Madabhushi, Parvin Mousavi, Septimiu Salcudean, James Duncan, Tanveer Syeda-Mahmood, and Russell Taylor, editors, Medical Image Computing and Computer Assisted Intervention – MICCAI 2023, pages 420–428, Cham, 2023. Springer Nature Switzerland. ISBN 978-3-031-43993-3.

Tom F Sterkenburg and Peter D Grünwald. The no-free-lunch theorems of supervised learning. Synthese, 199(3): 9979–10015, 2021.

Hugo Touvron, Thibaut Lavril, Gautier Izacard, Xavier Martinet, Marie-Anne Lachaux, Timothée Lacroix, Baptiste Rozière, Naman Goyal, Eric Hambro, Faisal Azhar, et al. Llama: Open and efficient foundation language models. arXiv preprint arXiv:2302.13971, 2023.

Nicholas J. Tustison, Brian B. Avants, Philip A. Cook, Yuanjie Zheng, Alexander Egan, Paul A. Yushkevich, and James C. Gee. N4itk: Improved n3 bias correction. IEEE Transactions on Medical Imaging, 29(6):1310–1320, 2010.

Pierre Vigier. perlin-numpy, 2020. URL https://github.com/pvigier/perlin-numpy.

Lennart Walger, Sophie Adler, Konrad Wagstyl, Leonie Henschel, Bastian David, Valeri Borger, Elke Hattingen, Hartmut Vatter, Christian E. Elger, Torsten Baldeweg, Felix Rosenow, Horst Urbach, Albert Becker, Alexander Radbruch, Rainer Surges, Martin Reuter, Fernando Cendes, Zhong Irene Wang, Hans-Jürgen Huppertz, and Theodor Rüber. Artificial intelligence for the detection of focal cortical dysplasia: Challenges in translating algorithms into clinical practice. Epilepsia, 64(5):1093–1112, 2023. URL https://onlinelibrary.wiley.com/doi/abs/10.1111/epi.17522.

Alan Q Wang, Batuhan K Karaman, Heejong Kim, Jacob Rosenthal, Rachit Saluja, Sean I Young, and Mert R Sabuncu. A framework for interpretability in machine learning for medical imaging. IEEE Access, 12:53277–53292, 2024a.

Alan Q Wang, Rachit Saluja, Heejong Kim, Xinzi He, Adrian Dalca, and Mert R Sabuncu. Brainmorph: A foundational keypoint model for robust and flexible brain mri registration. arXiv preprint arXiv:2405.14019, 2024b.

Kevin Y. Wang, Oluwatoyin R. Idowu, and Doris D.M. Lin. Chapter 24 - radiology and imaging for cavernous malformations. In Robert F. Spetzler, Karam Moon, and Rami O. Almefty, editors, Arteriovenous and Cavernous Malformations, volume 143 of Handbook of Clinical Neurology, pages 249–266. Elsevier, 2017. URL https://www.sciencedirect.com/science/article/pii/B9780444636409000242.

Jörg Wellmer, Carlos M. Quesada, Lars Rothe, Christian E. Elger, Christian G. Bien, and Horst Urbach. Proposal for a magnetic resonance imaging protocol for the detection of epileptogenic lesions at early outpatient stages. Epilepsia, 54(11):1977–1987, 2013. URL https://onlinelibrary.wiley.com/doi/abs/10.1111/epi.12375.

David H Wolpert. The lack of a priori distinctions between learning algorithms. Neural computation, 8(7):1341–1390, 1996.

Julian Wyatt, Adam Leach, Sebastian M. Schmon, and Chris G. Willcocks. Anoddpm: Anomaly detection with denoising diffusion probabilistic models using simplex noise. In 2022 IEEE/CVF Conference on Computer Vision and Pattern Recognition Workshops (CVPRW), pages 649–655, 2022.

Ziv Yaniv, Bradley C. Lowekamp, Hans J. Johnson, and Richard Beare. SimpleITK image-analysis notebooks: a collaborative environment for education and reproducible research. Journal of Digital Imaging, 31(3):290–303, Jun 2018. ISSN 1618-727X. URL 10.1007/s10278-017-0037-8.

